# Coronavirus Disease 2019 (COVID-19): An Evidence Map of Medical Literature

**DOI:** 10.1101/2020.05.07.20093674

**Authors:** Nan Liu, Marcel Lucas Chee, Chenglin Niu, Pin Pin Pek, Fahad Javaid Siddiqui, John Pastor Ansah, David Bruce Matchar, Sean Shao Wei Lam, Hairil Rizal Abdullah, Angelique Chan, Rahul Malhotra, Nicholas Graves, Mariko Siyue Koh, Sungwon Yoon, Andrew Fu Wah Ho, Daniel Shu Wei Ting, Jenny Guek Hong Low, Marcus Eng Hock Ong

**Affiliations:** Duke-NUS Medical School, National University of Singapore, Singapore; Health Service Research Centre, Singapore Health Services, Singapore; Faculty of Medicine, Nursing and Health Sciences, Monash University, Melbourne, Australia; Department of Emergency Medicine, Singapore General Hospital, Singapore; Department of Medicine, Duke University School of Medicine, Durham, NC, USA; Department of Anaesthesiology, Singapore General Hospital, Singapore; Department of Respiratory and Critical Care Medicine, Singapore General Hospital, Singapore; Singapore National Eye Centre, Singapore; Department of Infectious Diseases, Singapore General Hospital, Singapore

## Abstract

Since the beginning of the COVID-19 outbreak in December 2019, a substantial body of COVID-19 medical literature has been generated. As of May 2020, gaps in the existing literature remain unidentified and, hence, unaddressed. In this paper, we summarise the medical literature on COVID-19 between 1 January and 24 March 2020 using evidence maps and bibliometric analysis in order to systematically identify gaps and propose areas for valuable future research. The examined COVID-19 medical literature originated primarily from Asia and focussed mainly on clinical features and diagnosis of the disease. Many areas of potential research remain underexplored, such as mental health research, the use of novel technologies and artificial intelligence, research on the pathophysiology of COVID-19 within different body systems, and research on indirect effects of COVID-19 on the care of non-COVID-19 patients. Research collaboration at the international level was limited although improvements may aid global containment efforts.

## Introduction

On 11 March 2020, the Director-General of the World Health Organization (WHO), Dr Tedros Adhanom Ghebreyesus, declared Coronavirus disease 2019 (COVID-19) a pandemic, approximately 11 weeks after the first detected case of pneumonia of unknown aetiology in Wuhan, China was reported to the WHO Country Office in China on 31 December 2019.^1^ As of 6 May 2020, 3,623,803 cases of COVID-19 have been reported in 209 countries and territories, including 256,880 deaths.^2^ COVID-19 is caused by the novel betacoronavirus SARS-CoV-2, which is genetically similar to but distinct^3^ from other coronaviruses responsible for global outbreaks such as SARS-CoV-1^4,5^ and MERS-CoV.^6^

COVID-19 has attracted tremendous interest from researchers and clinicians worldwide, resulting in an appreciable body of COVID-19 literature being published in a relatively short period. Given the urgent need for evidence to support clinical and public health decisions, researchers have begun summarising and analysing the published literature to aggregate current evidence in the form of systematic reviews^7-9^ and bibliometric analyses.^10-12^ Existing bibliometric analyses^11,12^ have provided overviews of the COVID-19 research landscape. However, they primarily focus on authorship, keyword, and collaboration patterns without identifying gaps in the literature. More recent work includes a living mapping and living systematic review of randomised controlled trials (RCTs) which provides an up-to-date overview of the highest quality evidence on the prevention and treatment of COVID-19.^13^ However, to date, there has been no investigations of current gaps and future directions in COVID-19 research of all study designs and review types.

In this paper, we summarised the medical literature on COVID-19 between 1 January and 24 March 2020 using evidence maps and bibliometric analysis in order to systematically identify gaps and propose areas for valuable future research. Evidence maps in this study were a variant of the Evidence Gap Map (EGM), which is a systematic approach to identifying and describing the research activity in a topic area or policy domain, often through a focussed study of systematic reviews.^14,15^ EGMs have been used in a variety of research domains to characterise topic distributions and to inform priority setting in future research.^15,16^

## Methods

### Search strategy and selection criteria

We searched PubMed and Embase databases from 1 January to 24 March 2020 for the keywords “COVID” or “coronavirus” in the title or abstract. We used only these two terms to conduct a broad search that would ensure inclusion of the relevant literature. The search period was chosen on the premise that all articles on COVID-19 were published after the first report of the disease from the Wuhan government on 31 December 2019.^1^ We included all English language, COVID-19 related scientific articles, reviews, clinical case reports and series, and excluded duplicate articles, editorials, news, commentaries, and opinion pieces.

### Literature selection and data extraction

All extracted literature entries were exported into Microsoft Excel for screening and selection. Between 25 March 2020 and 7 April 2020, four reviewers (NL, MLC, CN, and PPP) independently screened the titles, abstracts, and, if ambiguous, full texts for the inclusion of articles. Discrepancies were resolved through discussions among the four reviewers, and in consultation with a fifth reviewer (FJS), to reach a consensus.

Subsequently, NL, MLC, CN, and PPP independently conducted information extraction from the included literature. Discrepancies were similarly resolved through discussion among the reviewers and in consultation with FJS.

### Bibliometric analysis

We retrieved basic bibliometric information from online full-text articles of all included articles, including publication date, number of days from submission to online publication, country of the first affiliated institution of the first author, number of countries and institution represented by the co-authors, and number of co-authors. Citation counts were retrieved from Google Scholar on 7 April 2020. Missing data for the number of days from submission to online publication was common and was treated using pairwise deletion. Other forms of missing data were excluded from the study using listwise deletion.

We performed bibliometric analysis on the retrieved information and presented the results as median (interquartile range [IQR]), proportions, rankings, or other descriptive statistics where appropriate. Additionally, we used the Hersch index (h-index) to measure the combination of quality and quantity of research output. An entity (i.e. country or author) has an h-index of *h* when it has a maximum of *h* articles with at least *h* citations.^17^ Trends in collaboration were analysed using cross-tabulation of international and inter-institutional collaboration; we reported the number (%) of articles that had both international and inter-institutional collaboration, only inter-institutional collaboration, and no international or inter-institutional collaboration.

All bibliometric analyses were conducted using Python version 3.8.0 (Python Software Foundation, Delaware, USA). Data of confirmed COVID-19 cases was obtained from the OurWorldInData.org dataset,^18^ which aggregates information from daily statistics published by the European Centre for Disease Prevention and Control.^2^

### Evidence map analysis

To generate evidence maps, we extracted additional information on the type, topic, and medical speciality of the articles. We categorised original articles into one of the following types: “Observational Research”, “Interventional Research”, “Protocol”, “Research” (basic science research and mathematical or computerised modelling works) or “Case Reports/Series”. Review articles, including narrative and systematic reviews and guidelines, were categorised as “Review”.

Based on the primary focus of the articles, we classified them into one of the following topics: “Basic Science” (articles on basic science or -omics research), “Epidemiology” (including patient risk factors, epidemiological characteristics, and disease trajectory), “Clinical Features and Diagnosis” (articles on all diagnostic elements such as patient signs and symptoms and radiologic or laboratory findings, including molecular diagnosis), “Pathogenesis” (articles reporting on viral mechanisms and disease progression, including immunology), “Treatment” (articles reporting all forms of management, including vaccines), “Public Health” (articles on public health and public policy), “Media” (articles reporting social media related topics), “Technology” (articles reporting the use of technology such as artificial intelligence and smart hardware), and “Health Economics” (articles reporting cost-benefit analysis, quality of life, and related topics). Articles with broad coverage of topics or without a singular focus were classified as “Overview” articles. The topics covered within each overview article were recorded and the distribution of articles covering each topic was presented.

We further classified articles into pre-determined medical specialities. As all articles on COVID-19 are related to infectious and respiratory diseases in some capacity, non-multidisciplinary articles were classified as “Infectious and Respiratory Diseases”, including respiratory medicine and internal medicine as contents of these articles were similar. Multidisciplinary articles were categorised by their primary speciality into one of the following: “Emergency and Prehospital Care”, “Critical Care” (including intensive care), “Anaesthesiology”, “Ophthalmology”, “Dentistry”, “Cardiology”, “Gastroenterology” (including hepatology), “Nephrology”, “Radiology”, “Pathology”, “Oncology”, “Paediatrics”, “Obstetrics and Gynaecology”, “Psychiatry” (inclusive of mental health-related articles), “Preventive Medicine” (inclusive of public health), and “Family Medicine” (inclusive of general practice-related articles).

To summarize the landscape of current COVID-19 research, several tables were created based on the cross-tabulation of article topic with article types, specialities, and continent of origin. The proportion of papers of a certain topic between two continents was compared using the chi-square test. Statistical significance was set at p<0.05.

## Results

The search between 1 January and 24 March 2020 yielded 1703 articles, of which 550 articles (32·3%) met the inclusion criteria for analysis. Figure 1 illustrates the details of the selection process.

**Figure 1:**
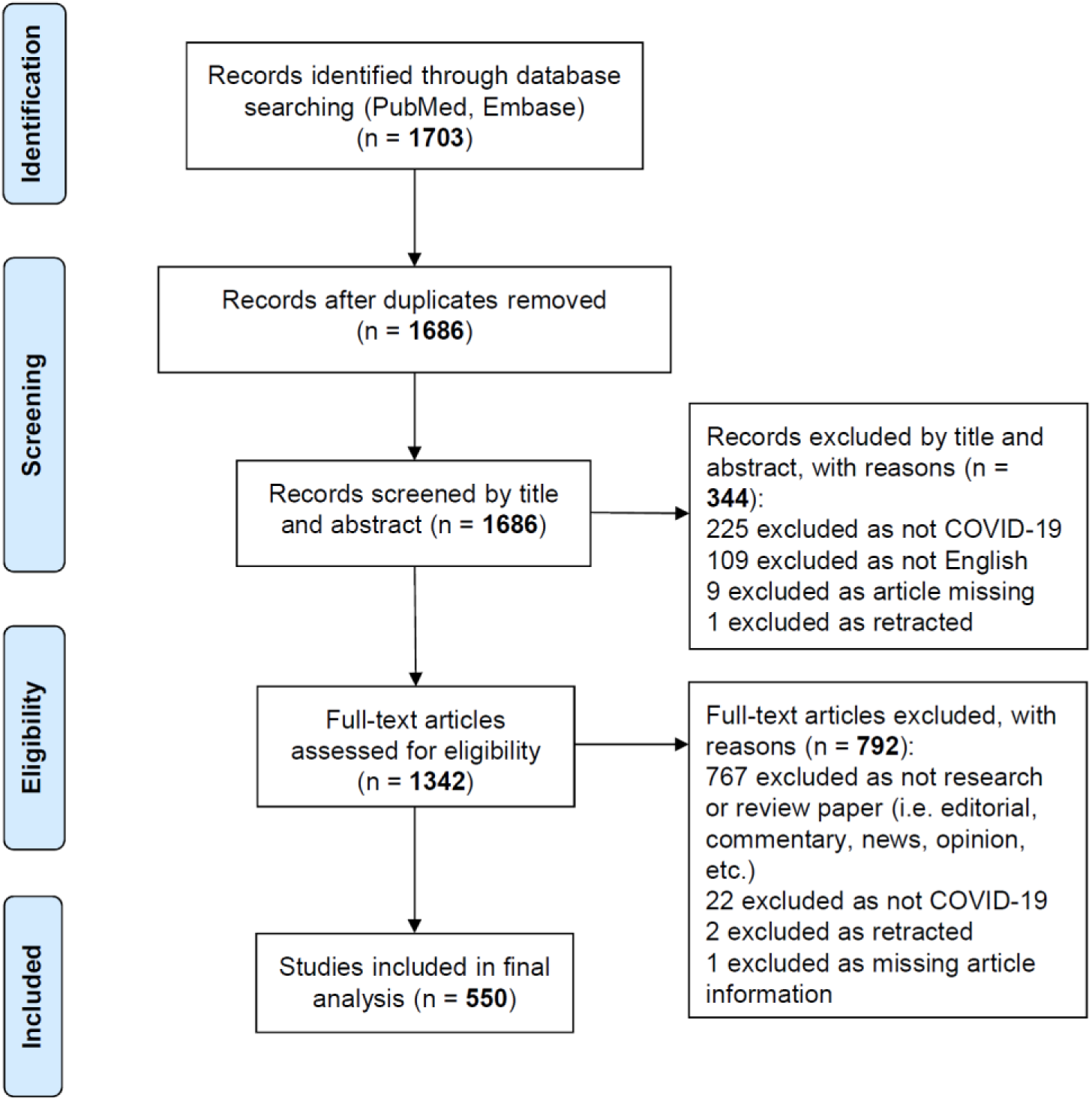
PRISMA flow diagram.

### Bibliometric features

Articles were published online over a period of exactly ten weeks from 14 January 2020 to 23 March 2020, with no included articles published on 24 March. The number of new articles per week increased steadily over the first nine weeks from one in the first week to 119 in the ninth week, dropping to 75 new articles in the final week from 17 to 23 March. Articles for which publication time data was available (*n*=373, 67·8%) had a median duration of eight days from submission to online publication (IQR: 4-16). The *h*-index for all 550 included articles was 57, with a cumulative total of 17450 citations.

First authors of the articles were from 33 countries (Figure 2). China produced the highest output with 323 articles (*h*-index=48), followed by the United States of America (USA) with 59 articles (*h*-index=16). However, 18 of the 33 countries (54·5%) published three or fewer articles.

**Figure 2:**
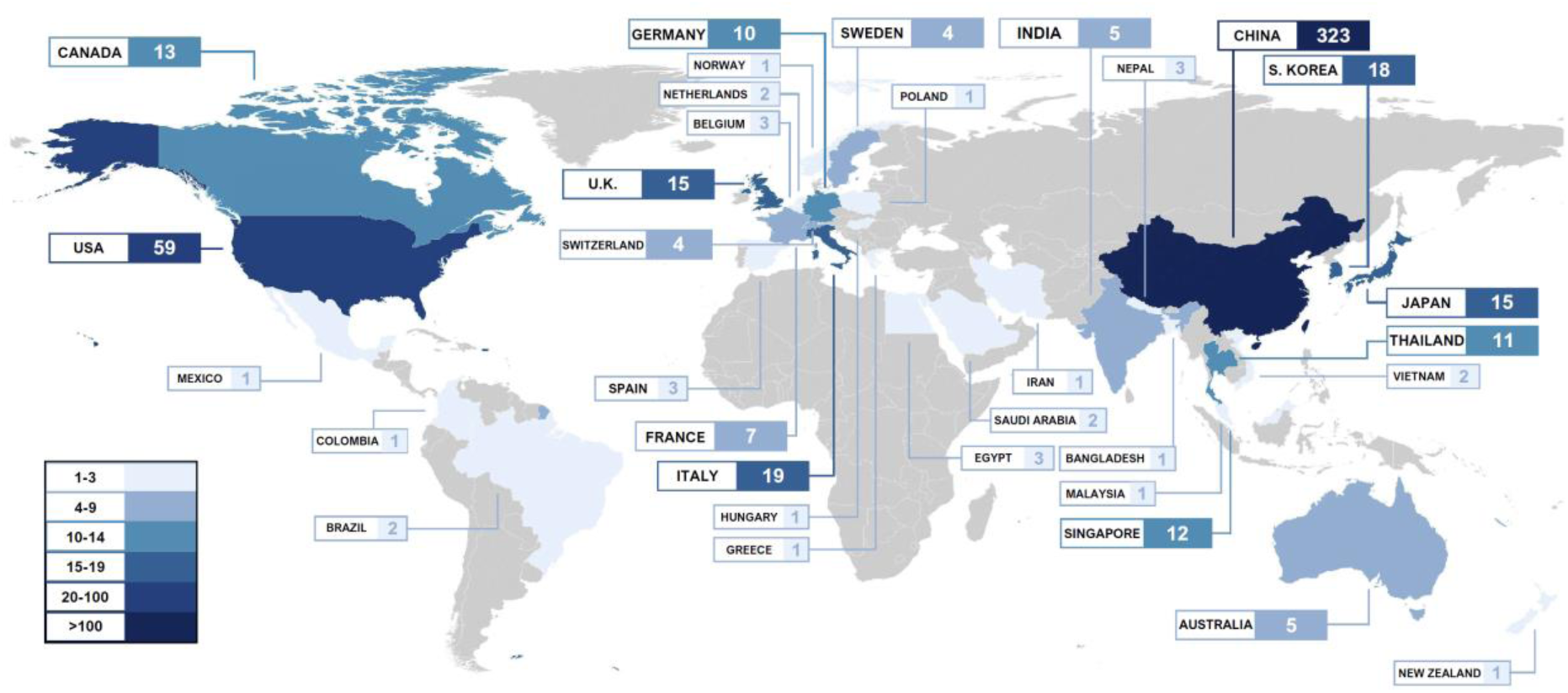
Distribution of articles by country.

As of 24 March 2020, articles from Asia (*n*=394), North America (*n*=73), and Europe (*n*=71) represented 71·6%, 13·3%, and 12·9% of all articles (Figure 3(a)), respectively, while confirmed COVID-19 cases from these same continents represented 33·6%, 13·1%, and 51·0% of 377261 global cases (Figure 3(b)), respectively.

**Figure 3:**
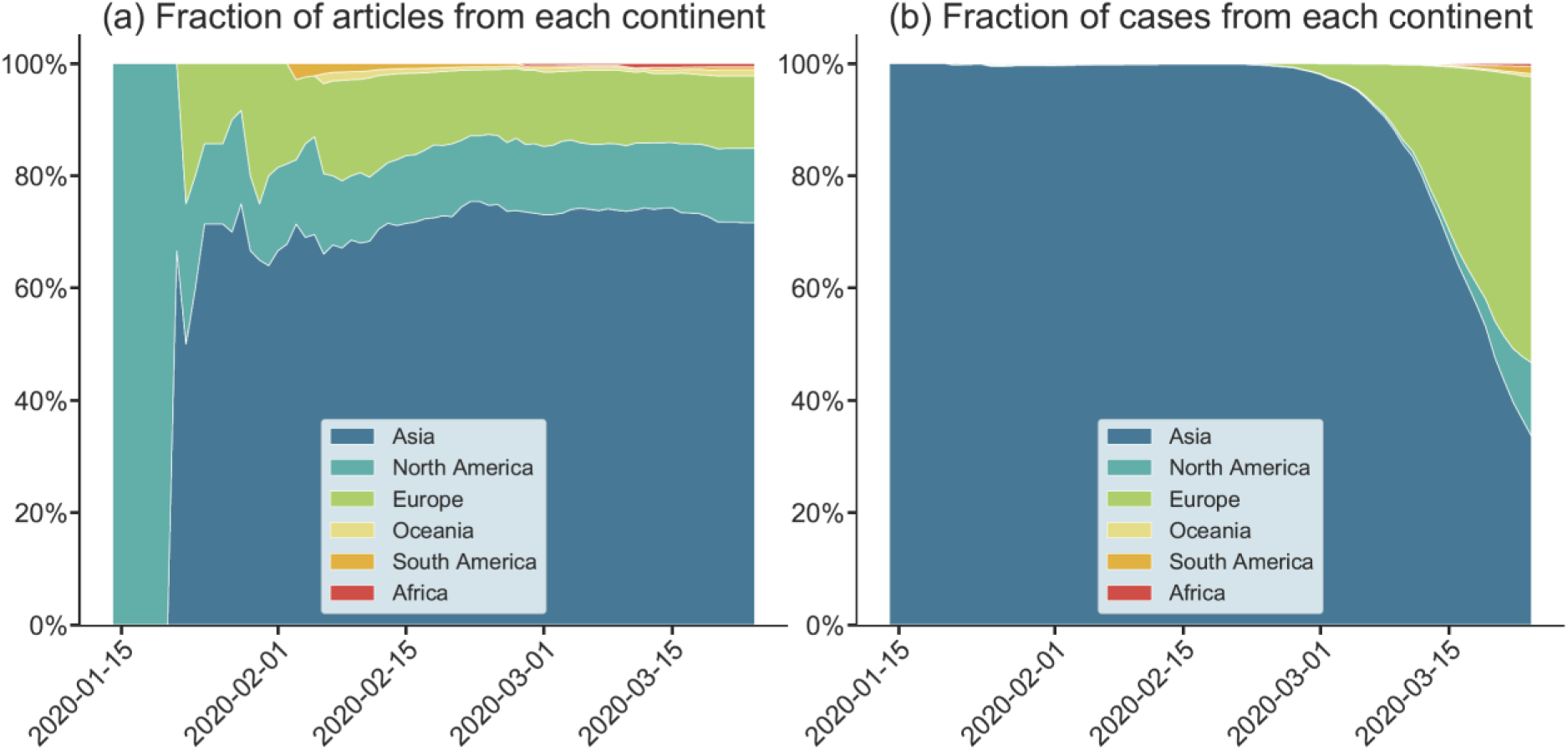
Fractions of (a) articles and (b) cases from each continent from 14 January to 24 March 2020.

Overall, 183 (33·3%) articles were written by authors from a single institution in one country, 231 (42·0%) articles were written by authors from multiple institutions within one country, while only 136 (24·7%) articles had collaboration between multiple institutions from multiple countries. Articles had a median of six co-authors (IQR: 4-10), including the first author and regardless of institution or country.

### Evidence maps

Figures 4 and 5 show the distributions of article topics by article type, continent of origin, and clinical speciality.

**Figure 4:**
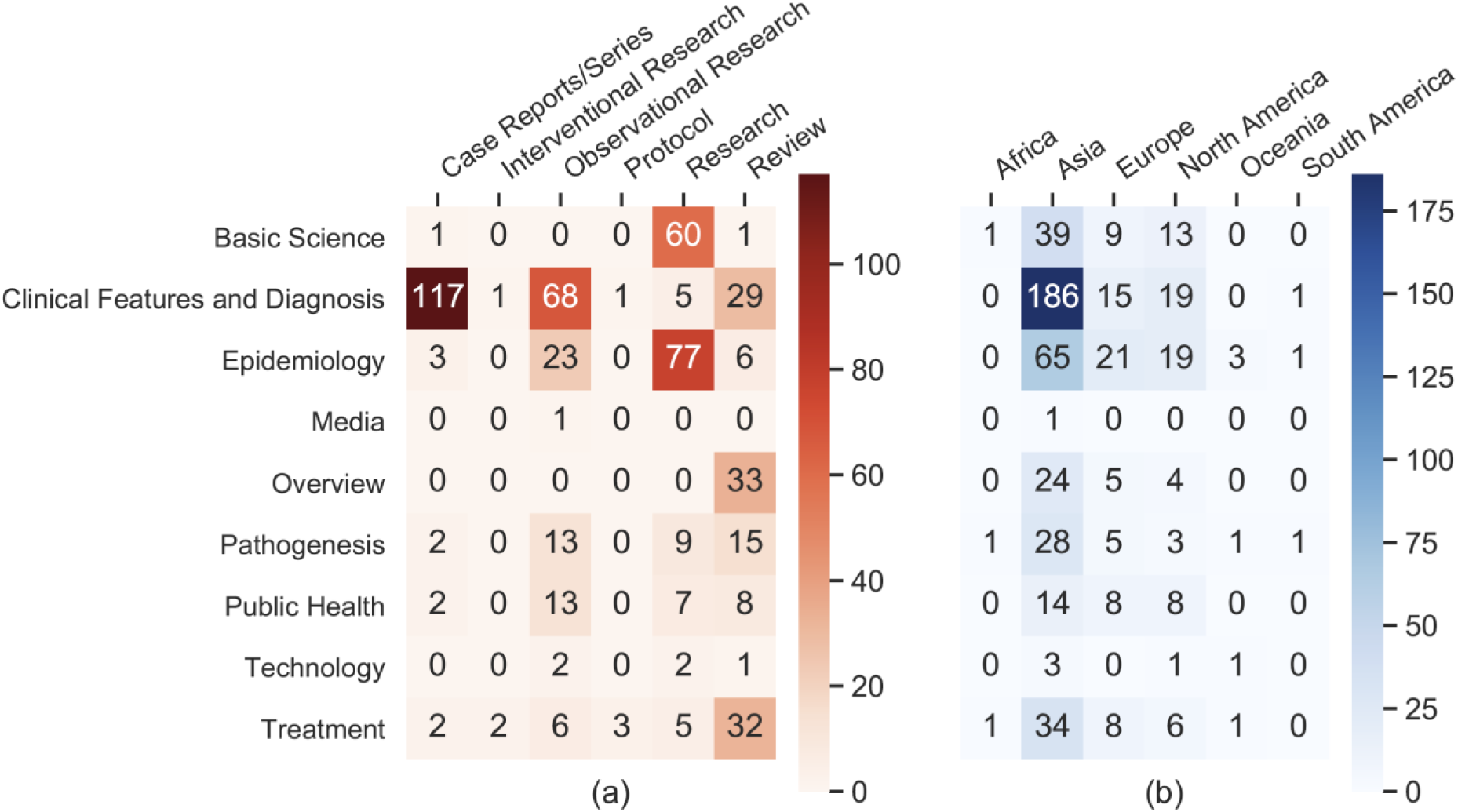
Evidence maps of the distribution of article topics by (a) article type and (b) continent of origin.

**Figure 5:**
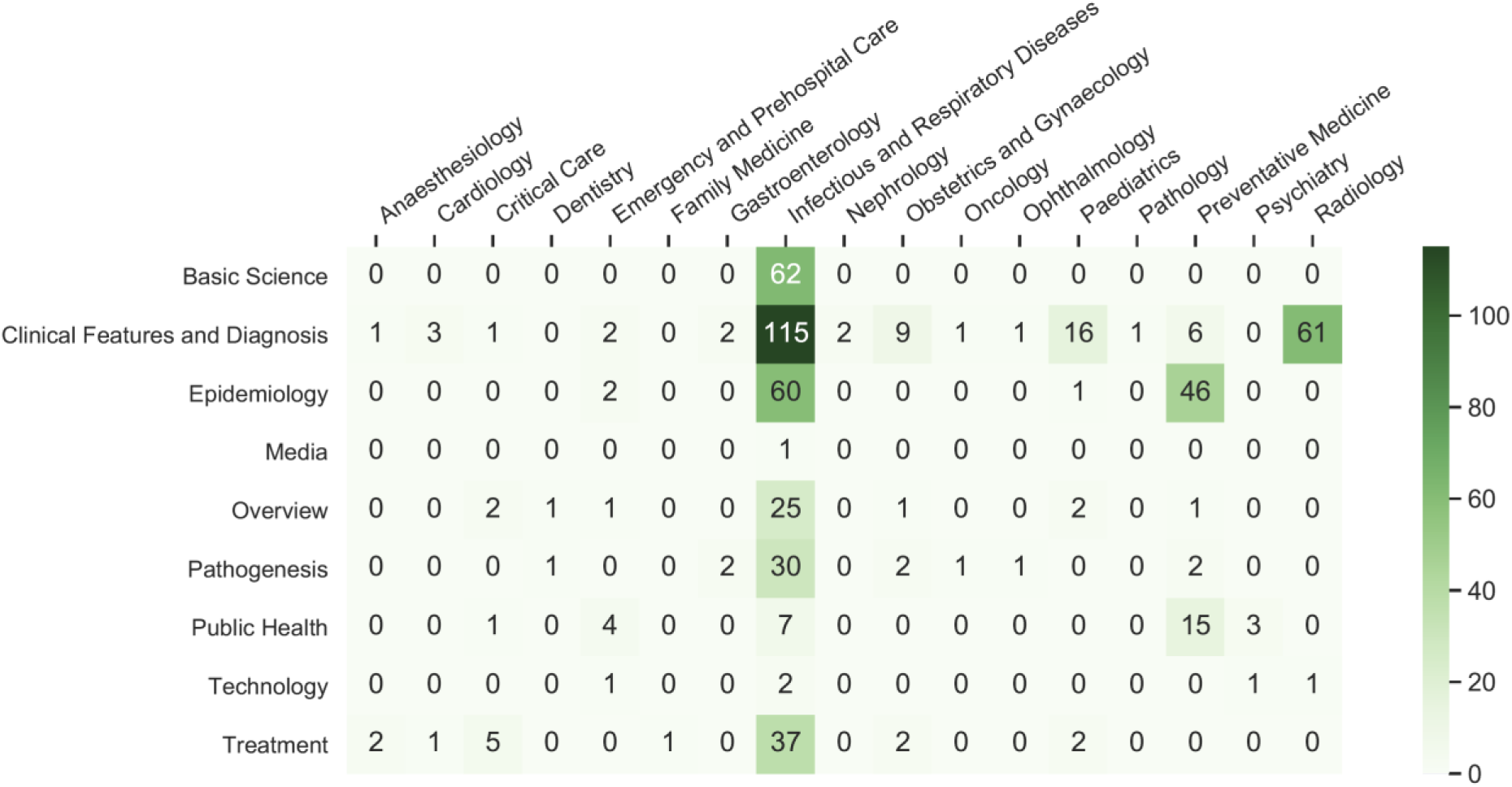
Evidence map of the distribution of article topics by medical speciality.

According to article topic alone, clinical presentation and diagnosis was the most common (40·2%), followed by epidemiology (19·8%), basic science (11·3%), treatment (9·1%), pathogenicity (7·1%), overview (6·0%), public health (5·5%), technology (0·9%), and media (0·2%). No articles focussed on health economics. By article type alone, general research papers comprising mainly epidemiological or virologic studies were the most common (30·0%), followed by case reports/series (23·1%), observational research (22·9%) and review papers (22·7%). There were few articles on trial protocols (*n*=4, 0·7%) and interventional research (*n*=3, 0·5%), of which only one study was conducted *in vivo*.

By article topic and type (Figure 4(a)), case reports/series on clinical features and diagnosis (*n*=117, 21·2%) were the most common, followed by general epidemiological research (*n*=77, 14·0%) comprising mainly of studies that modelled disease trajectory. Observational research on clinical features and diagnosis (*n*=68, 12·4%) and general research on basic science (*n*=60, 10·9%) were also common. There were 33 review papers classified as overview papers, which were further analysed to elucidate the topics covered (Figure 6). Most overview papers shared similar topics, including clinical features and diagnosis, epidemiology, treatment, pathogenicity, and public health. Only five out of 33 overview papers reported on basic science.

**Figure 6:**
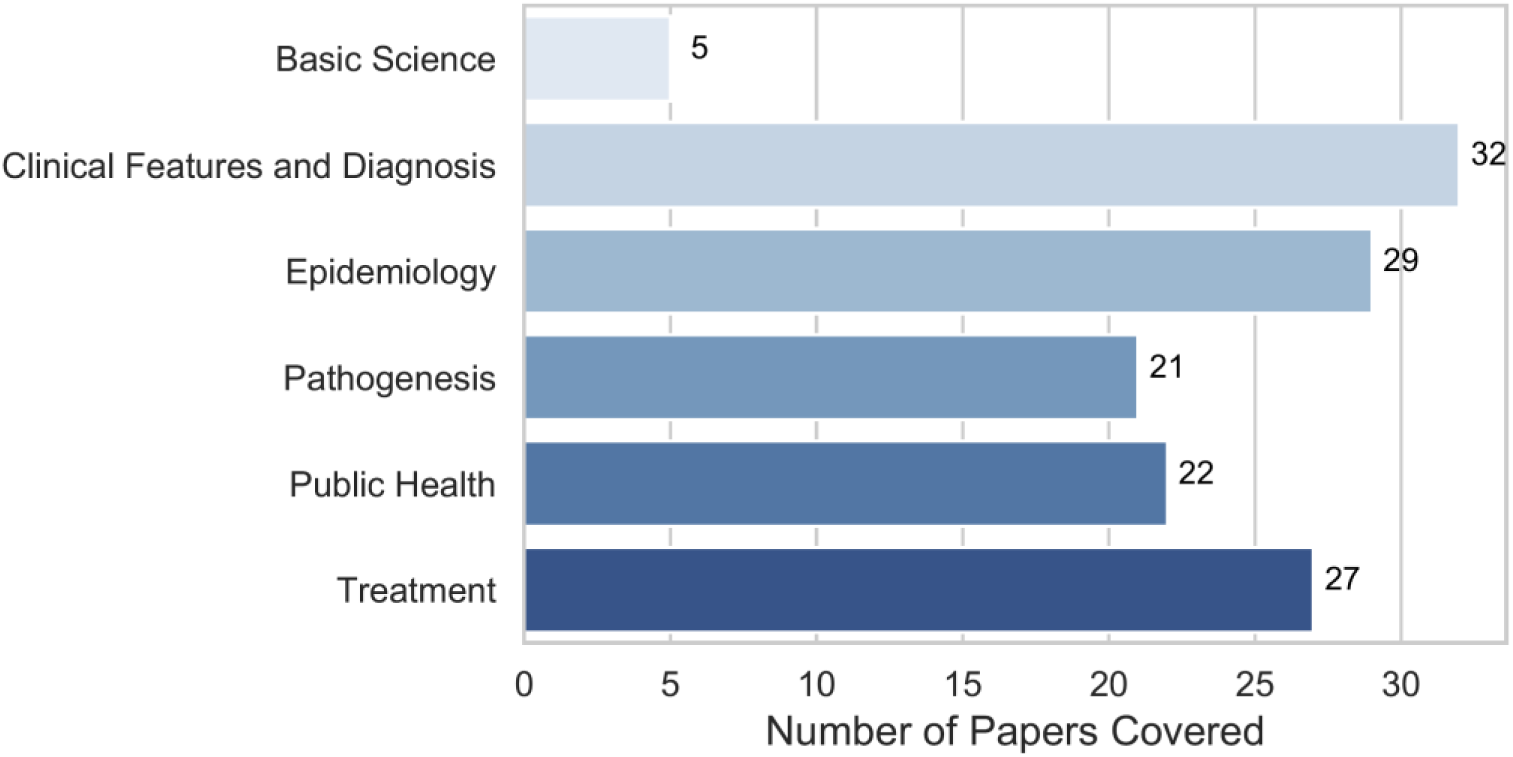
Distribution of topics of overview papers.

According to the evidence map in Figure 4(b) which analysed article topic and continent of origin, papers on clinical features and diagnosis published in Asia were the most common (*n*=186, 33·8%). Moreover, the proportion of articles reporting on clinical features and diagnosis within articles from Asia (186/394, 47·2%) is significantly higher than the proportion of such articles from North America (19/73, 26·0%), and Europe (15/71, 21·1%), with both p-values being less than 0·01. However, for the topic of epidemiology, the proportions of articles from Europe (21/71, 29·6%, p=0·01) and North America (19/73, 26·0%, p=0·07) were higher compared to Asia (65/394, 16·5%).

In terms of the primary speciality of articles, articles on infectious and respiratory diseases predominated (*n*=61·6%), followed by articles with primary specialities of preventive medicine (*n*=70, 12·7%), radiology (*n*=62, 11·2%), and paediatrics (*n*=21, 3·8%). As shown in Figure 5, apart from articles on infectious and respiratory diseases, articles on radiological features and diagnosis were the most common (*n*=61, 11·1%), followed by preventive medicine articles on epidemiology (*n*=46, 8·4%) and public policy (*n*=15, 2·7%) and articles on clinical features and diagnosis in paediatric patients (*n*=16, 2·7%) and obstetric and gynaecological patients (*n*=9, 1·6%). All other specialities not reported above were lacking in articles, having three or fewer articles on clinical features and diagnosis and, apart from critical care, two or fewer articles on treatment.

## Discussions

Since the outbreak of COVID-19 in December 2019, more than 1200 relevant articles (as of 24 March 2020) have been published in scientific journals, of which 767 were editorials, commentaries, news, and opinions. Additionally, as of 6 May 2020, 2830 preprints have been archived in medRxiv (2255) and bioRxiv (575) servers. COVID-19 has garnered research interest faster than any other pandemic in history, possibly due to its high transmissibility^19^ fuelled by global interconnectivity; there were fewer articles on SARS and MERS combined within a year after their initial outbreaks than COVID-19 articles within the first three months of its discovery.^20,21^ Not surprisingly, one of the first reports^22^ on clinical features of patients infected with COVID-19 had attracted 1411 citations within two months of publication; its citations reached 3845 by 6 May 2020.

While there was a large number of articles on epidemiological characteristics and clinical features and diagnosis of COVID-19 patients, there was a dearth of papers within our search period that reported findings from RCTs on drugs and treatments; only one clinical trial was conducted on human subjects.^23^ This is likely due to our search period ending on 24 March 2020, which is considered early in the pandemic. As such, there was insufficient time to design, approve, and execute such trials. Furthermore, we did not search clinical registries such as ClinicalTrials.gov, which indexes clinical trials globally, as the focus of our study was published literature. From the time since our analysis, we observed an increased number of clinical trials and protocols being published and summarised.^13^

Another observation is the paucity of technology-related articles in the COVID-19 medical literature. While digital technologies such as artificial intelligence (AI), big data, and the internet have positively impacted public health intervention strategies^24^ and shown promising applications in infectious disease contexts,^25^ few have looked into their applications in COVID-19 research. Despite the straightforward application of AI technology (in particular, deep learning)^26^ in analysing medical images, there was only one such study on chest computed tomography (CT) within articles on radiology.^27^ Furthermore, despite the high volume of articles on epidemiology, current literature mainly adopted traditional techniques^28^ to analyse COVID-19 dynamics^29-31^ and forecast trends in the COVID-19 pandemic^32^ and its trajectory.^33^ Traditional statistical methods rely heavily on underlying assumptions which do not apply to medical data which are multidimensional, dynamic, and highly nonlinear as unpredictable human-environment interactions are involved.^25^ More robust and complex modelling has many promising public health applications in COVID-19,^34^ with similar methods already validated in tuberculosis and gonorrhoea epidemic control^35^ and malaria policy.^36^ The utility of such models extends to lesser investigated areas in COVID-19 research including social media and public reaction analysis.^37,38^

As observed in our evidence maps on topic and speciality, apart from articles which focussed primarily on infectious and respiratory diseases, there have been substantial works in preventive medicine and radiology, as well as some articles from population-specific specialities such as paediatrics and obstetrics and gynaecology. However, few articles are seen in all other specialities, leaving obvious gaps in research. For instance, acute kidney and cardiac injury were among the top adverse clinical complications observed in COVID-19 patients with severe disease,^22^ which should prompt more research on the pathophysiology of disease within specialities like cardiology, nephrology, and critical care where research is currently sparse. Furthermore, with community clinics, emergency medical services, and emergency departments experiencing a surge in patients, research into the efficient allocation of resources, such as adaptive bed management and operation scheduling, is much needed.

Health care workers have been confronted with unprecedented levels of morbidity and mortality, as well as the constant threat of being exposed to the virus. Quarantine, a common public health measure globally in this pandemic, has significant psychological impacts on those affected.^39^ Also, COVID-19 patients and their families can face undue stress as a result of self-blame, fear of transmitting the virus, and uncertainties regarding their health. These are just a few of a multitude of factors that contribute to high psychological stress in persons with and without COVID-19. Regrettably, one of the first research articles exploring the mental health of health care workers in this pandemic was only published at the end of our study’s inclusion period.^40^ While other viewpoints,^41,42^ guidelines,^43^ and research^44^ on mental health have subsequently surfaced, mental health and psychiatry in all populations remain grossly underrepresented even within the latest literature. Other major clinical settings to explore include but are not limited to neurology,^45^ anaesthesiology,^46^ cancer,^47^ pathology, and geriatrics. It is also important to note that the impact of this pandemic extends beyond COVID-19 patients,^48^ highlighting the importance of providing high-quality, equal, and continuous care to non-COVID-19 patients. Research should thus be done to quantify the severity and extent to which medical and social care for patients with subacute or chronic conditions have been affected, as well as to uncover other unintended consequences on non-COVID-19 patient care.

Our analysis revealed that the submission-to-publication time for COVID-19 articles was much shorter than normal, indicating an accelerated peer-review process. Numerous journals have prioritised COVID-19-related research, giving clinicians, researchers, and the public quicker access to peer-reviewed articles. While this phenomenon is encouraging and a testament to the swift response of the scientific community, acceleration of the peer-review process can potentially lead to cursory reviewing and lax publication standards from journals and hasty publication by authors; consequently, the integrity of research may be compromised.^49^ The undesirable consequences associated with accelerated publication have already surfaced in recent, potentially misleading studies which reported on the positive effects of hydroxychloroquine in COVID-19.^50-52^ Despite the poor study design of these articles^53-55^ and potential cardiotoxicity of hydroxychloroquine,^56^ there was an increase in public demand for hydroxychloroquine following endorsements of its use in COVID-19 treatment.^57^ Editors, reviewers, and authors alike have the responsibility of maintaining the integrity of the peer-review process, despite the need for accelerated publication during this crucial period.

With the increasing proportion of global cases in Europe and North America both during and after our period of analysis, the predominance of articles originating from Asia — particularly articles on clinical features and diagnosis — necessitates additional research and evidence from these continents. In the period after our analysis, we observed an increasing number of new articles and preprints from continents other than Asia, although this trend was not captured in our analysis. Cross-country collaborations and initiatives have been limited based on our analysis, despite their essential role in facilitating multi-site clinical trials and patient data sharing. A heartening example is seen in the formation of an international consortium (4CE) consisting of 96 hospitals across 5 countries^58^, in which harmonized electronic health record (EHR) data were analysed locally and converted to a shared aggregate form to analyse and visualise regional differences and global commonalities.

With increased understanding of COVID-19, new challenges have also emerged. Unlike SARS, many COVID-19 patients are mildly symptomatic or even asymptomatic^59^, which makes screening and identification of cases extremely difficult and leaves the public at risk of infection. The gold standard test based on reverse transcriptase-polymerase chain reaction (RT-PCR) is accurate but is time-consuming, while alternative rapid test kits are fast and make point-of-care diagnosis possible but have shown disappointing performance to date. In general, the current diagnostic tools and technologies are not readily available for large-scale, population-based screening of COVID-19, leaving much room for future research.

Besides those discussed, there are still many unknowns that will require investments from both public and private entities to resolve. For instance, to what extent will a vaccine slow the spread COVID-19? Are repeated infections possible and, if so, how long after the first infection? What is the quality of life among survivors with severe disease? What are effective public health measures at the national and international levels that can retard SARS-CoV-2 transmission while minimising the impact on global citizens and the global economy? Are current infection control measures in prehospital and hospital settings sufficient and, if not, how can they be improved? We should note that the damage of COVID-19 has propagated far beyond the healthcare sector into almost all other industries. Furthermore, while conditions in some countries are beginning to ease, conditions in others are deteriorating. With the evidence maps investigated in our study, we hope to unveil some of the critical gaps and blind spots within the initial body of COVID-19 research and provide meaningful direction for future research.

### Limitations

There are several limitations to our study. Firstly, our search period did not fully encapsulate the period in which the majority of COVID-19 cases shifted from China to Europe and the USA. Therefore, the results do not fully describe the most recent research landscape due to the exclusion of newly published articles. Furthermore, we did not search other databases such as ClinicalTrials.gov, which would have excluded most clinical trial protocols. Also, only English language articles were analysed which resulted in the exclusion of articles from China — a substantial source of early COVID-19 literature. Lastly, the exclusion of non-peer reviewed research (those archived in medRxiv and bioRxiv) in our analysis may have neglected some new evidence but ensured the inclusion of only scientific results that have undergone peer review.

## Conclusions

COVID-19 research is only in its nascent phase, but a large volume of research articles has already been published. However, there are still knowledge gaps in the literature that have yet to be filled. Of particularly concern and urgency are the effects of COVID-19 on the mental health of health care workers, patients, and other populations. Other underexplored areas of research include the pathophysiology of COVID-19 within different body systems and populations, the use of novel technologies such as artificial intelligence in forecasting trends and improving prevention and intervention strategies, and the indirect effects of the pandemic on the care of non-COVID-19 patients. We opine that the systematic identification and filling of such gaps in the literature will be vital in informing and shaping effective, evidence-based governmental policies and healthcare practices, which can translate to improved outcomes.

## Data Availability

The datasets used and/or analyzed during the current study are available from the corresponding author on reasonable request.

## Contributors

NL conceived and designed the study. NL, MLC, CN, and PPP screened and reviewed the articles and extracted paper information, which were verified by FJS. NL, MLC, CN, PPP, and FJS planned the formal analyses and drafted the manuscript. MLC and CN analysed the data. All authors made substantial contributions to results interpretation and critical revision of the manuscript. All authors approved the final manuscript.

